# Estimating excess mortalities due to the COVID-19 pandemic in Lusaka, Ndola, and Kitwe urban districts in Zambia using burial records data

**DOI:** 10.64898/2025.12.25.25343016

**Authors:** Joseph Sichone, Musalula Sinkala, Sody M. Munsaka, Martin Simuunza

**Affiliations:** Department of Biomedical Sciences, School of Health Sciences, University of Zambia, Lusaka, Zambia; Department of Disease Control, School of Veterinary Medicine, University of Zambia, Lusaka, Zambia; Africa Centre of Excellence for Infectious Diseases of Humans and Animals, University of Zambia, Lusaka, Zambia

**Keywords:** COVID-19, excess mortalities, burial records, all-cause mortality, Zambia

## Abstract

**Introduction:** The true mortality impact of the COVID-19 pandemic is difficult to estimate at the national level due to limitations in testing, surveillance, and accurate attribution of cause of death. Therefore, developed nations have used estimates of excess mortality during the pandemic as an objective indicator based on historic national death registration data. This could not be estimated in developing African nations, including Zambia, due to incomplete death registration data hence the true impact of the pandemic in these regions remains potentially underestimated. Since the pandemic primarily concentrated in large cities, this study utilises burial records from Lusaka, Ndola, and Kitwe urban districts in Zambia to estimate COVID-19-associated excess mortalities.

**Methodology:** Linear and Negative Binomial Regression were applied on burial permits data, some dating back to 2011, to estimate monthly expected all-cause mortalities and corresponding excess mortalities during the 2020 - 2021 pandemic period. Pearson’s correlation and multivariate linear regression were also used to corroborate excess mortalities with COVID-19 incidence and other meteorological parameters.

**Results:** Kitwe and Ndola had substantial gaps in burial records while Lusaka had minimal gaps. On average, Kitwe recorded no excess mortalities while Lusaka and Ndola recorded significant excess mortalities with overall 3,484 (P-score 11.1%) and 378 (P-score 6.1%) excess deaths respectively. This translates to an estimated 3.5 for Lusaka and 1.5 for Ndola COVID-19 deaths undercount ratio. Excess mortality positively correlated with COVID-19 incidence (P < 0.05) suggesting that it was caused by the pandemic and negatively correlated with temperature after controlling for average relative humidity and average hours of sunshine (P < 0.05).

**Discussion and conclusion:** The findings of this study show that the mortality impact of COVID-19 was underestimated in Zambia and this may apply to other African nations. It further shows that pandemic excess mortalities can be tracked in African urban centres that have complete burial records data.

## Introduction

Despite its unrivalled global spread and devastating total death toll of over 7 million by May 2024 [1–4], the COVID-19 pandemic has shown variable intensity in the number of mortalities caused in different populations [5–8]. Although the reason for this is not fully known, it has been postulated that this disparity may, in part, be an outcome of limitations in testing, surveillance, and accurate attribution of cause of death among nations. Therefore, the true pandemic death toll may be higher worldwide [9–19]. This presents a challenge since accurate measurement of mortalities is vital for understanding the epidemiology of the disease and for guiding current and future public health policy [9,11,13,16].

The World Health Organisation (WHO) has therefore advised measuring excess mortality associated with the COVID-19 pandemic as an objective and comparable measure of the true mortality impact of the pandemic [11,20]. Excess mortality is the number of deaths from all causes during a public health crisis in a given place and time above what would have “normally” been expected based on historical norms [9,11,20–24]. In other words, it is the total observed deaths above the average expected all cause mortalities (ACM) for a particular period in a country. The ACM is projected using official national death registration data from national civil registration systems (CRS) spanning at least the previous five years in that population [9,11,16]. Excess mortalities are normally recorded either as absolute numbers or as “P-scores”; which is the percentage rise in total expected mortalities. Assuming there were no other causes of mass mortalities during the COVID-19 crisis such as wars or natural disasters, all excess mortalities observed during the pandemic can therefore be either directly or indirectly attributed to COVID-19. This gives us a better measure of the total mortality impact of the pandemic and also accounts for undocumented COVID-19 deaths [9,12,13,15]. Various early independent studies estimated that there were over a million excess mortalities within the first year alone of the COVID-19 pandemic (2020) across 29 different countries with officially reported COVID-19 deaths accounting for only between 20% to 78% of these excess mortalities [12,13,15,23]. In 2022, the WHO used mortality data from 2015 to 2021 and estimated that there were about 14.83 million excess deaths globally during the pandemic between 1^st^ January 2020 and 31^st^ December 2021 which means that there were 2.74 times more deaths than the 5.42 million reported as due to COVID-19 for the period [11]. The “Our World in Data” website [9] also holds an up-to-date database of COVID-19 associated excess mortalities for about 124 countries and territories worldwide estimated using all-cause mortality data from the World Mortality Dataset (WMD), Human Mortality Database (HMD), The Economist, and the World Health Organisation.

Unfortunately, COVID-19 excess mortalities could not be estimated for many countries especially in the Africa (87%), South East Asia (82%), Western Pacific (67%), Eastern Mediterranean (57%), and Americas (34%) WHO regions due to incomplete national death registration data in these countries [11]. In fact, as an example, according to the WHO and United Nations data, the completeness of death registration within the national CRS is as low as 2% in some African countries [38,39]. In Zambia, a developing nation in Sub-Saharan Africa, the completeness of official death registration by the national CRS was about 20% in 2017 [40] and only 18% in 2020 [43] nationally. For such nations lacking required data, only indirect estimates or approximations of excess mortalities can be obtained. For example, in their 2022 study mentioned above, the WHO used a Poisson count framework that applied Bayesian inference techniques and relied on ACM information from other countries for which data exists, as well as other relevant factors, to estimate excess mortalities for countries lacking data [11]. Their model therefore estimated that there were about 10% to 20% excess mortalities between January 2020 to December 2021 during the COVID-19 pandemic in Zambia as an example and this meant that COVID-19 deaths were likely 5 to 10 times more than what was officially reported in the country. However, reliance on such complex statistical models, though robust, may still have its limitations and may not be a replacement for the use of empirical data. An alternative and relatively more empirical indirect approach to estimating excess mortalities in such situations is to quantify changes in burial trends based on actual cemetery or burial permit data in the given place. In the COVID-19 era, this approach was first seen in a study by Elyazar and colleagues [41]. In their study, they estimated that there were about 61% excess mortalities in the first ten months of the COVID-19 pandemic in Jakarta, Indonesia, through a review of burial records data at public cemeteries in the city and demonstrated that this is an equally sensitive and timely method compared to the use of official death registration data at CRS departments in developed nations. One obvious feature of this approach is that it is best suited for estimating excess mortalities at the subnational level for smaller administrative units using local burial data. Therefore, the current study adopts this approach to estimate COVID-19 associated excess mortalities in Lusaka, Ndola, and Kitwe urban districts in Zambia which were among the top six most affected districts by the pandemic in Zambia and together contributed about 52% of all officially confirmed COVID-19 deaths in the whole country between 2020 – 2021 according to the Zambia National Public Health Institute (ZNPHI) data [42,43,72]. Further, since excess mortality is described as a more unbiased parameter for tracking COVID-19 disease impact than officially reported cases [9,11,16,20], this study also assessed the relationship between the estimated COVID-19 excess mortality and meteorological parameters such as temperature to help consolidate our understanding of COVID-19 disease dynamics and such parameters.

## Materials and Methods

### Study Areas

Lusaka district, also Lusaka city, is the capital of Zambia and the most urbanised and largest district by population in Zambia as well as the most densely populated in the country with a current population of just over 2.2 million and population density of about 5,270 people / square kilometre [45, 46, 47]. It is located in Lusaka province in the central part of the country and is the centre of commerce and government for Zambia [46, 47]. The first official cases of COVID-19 in Zambia were recorded in Lusaka district on 18^th^ March 2020 and, being the epicentre of the pandemic, the district had recorded about 63,527 confirmed cases and 990 confirmed COVID-19 deaths by December 2021 [42]. Lusaka district also has a very high mortality data capture rate due to a lack of village settlements in the district which are usually a source of incomplete mortality data reporting. For example, the mortality registration completeness by the CRS department for the year 2020 in Lusaka district was estimated to be higher than 80% even though this metric was lower at only 52.3% the same year for the entire Lusaka Province in which Lusaka district is located [44]. On the other hand, Kitwe is the second largest district in Zambia by population after Lusaka district with a current population of about 661,000 and population a density of about 814 people / square kilometre [45,47]. It is a developed commercial and industrial urban district located on the Copperbelt province of Zambia which is the major mining region of the country bordering the Democratic Republic of Congo (DRC) [48]. It is third only to Lusaka and Ndola districts in terms of Infrastructure development [48,49]. Between 2020 and 2021, Kitwe had recorded about 11,100 confirmed cases and 211 confirmed COVID-19 deaths [42]. Lastly, Ndola is the third largest district in Zambia by population after Lusaka and Kitwe districts with the latest population of about 624,000 and a population density of about 651.5 people / square kilometre [45]. It is a developed urban district also located on the Copperbelt province and is also the administrative capital of the province [49]. Between 2020 and 2021, Ndola had recorded about 12,220 confirmed cases and 250 COVID-19 deaths [42] but these numbers are thought to be magnitudes lower than the actual number of COVID-19 cases and deaths in all the districts due to poor surveillance and testing systems in the country [9,11,18,43]. Unfortunately, the official death registration completeness by the CRS department is low for the entire Copperbelt province at only about 35.7% in 2020 [44] and this is also expected to be reflective of the official CRS death registration rates for both Kitwe and Ndola districts due to the presence some village settlements in both these districts.

### Data Collection

Monthly all-cause mortality (ACM) data were collected in the form of the total number of burial permits issued per month by the local councils in Lusaka, Ndola, and Kitwe districts. This also included all available data on unclaimed bodies as well as “transfers”, which are deaths that occurred within the district but were transported to another district for burial. Flow chart 1 depicts this data source as step 3 in the general steps of official death registration by CRS in Zambia. Note that some steps in the flow chart occur concurrently [44,46].

**Flow chart 1.** General steps of official death registration with CRS in Zambia. Some steps in the flow chart occur concurrently.

Many deaths are not officially registered with the CRS department in step 5 partly due to logistical challenges or poor reporting due to the presence of village settlements in a particular district. However, a burial permit is normally always issued by the local council before any burial can take place at a public or private burial site in step 3. Therefore, step 3 (local council burial permits records) was chosen as the data collection point for this study with the expectation that it would hold more mortality information than the respective CRS departments in each study district. In Lusaka district, ACM (i.e. burial permits data) from January 2015 to December 2021 was therefore collected at the Lusaka city council offices located at the University Teaching Hospital morgue, Levy Mwanawasa University Teaching Hospital morgue, as well as at the Lusaka city council head office. This data was available for all six main burial sites in Lusaka district namely “Leopards Hill”, “Chingwere”, “Chunga”, “Memorial Park”, “Mutumbi”, and “Kabanana”. In the Ndola district, similar data from January 2011 to December 2021 was collected at the Ndola city council head office. This included data for all four main burial sites in the district owned by the council namely “Kansenshi”, “Kawama”, “Mitengo”, and “Kantolomba”. Lastly, in Kitwe district mortality data from January 2015 to December 2021 were collected at the Kitwe city council office located at the Kitwe Teaching Hospital morgue as well as the Kitwe city council head office. This included data for all six main burial sites owned by Kitwe city council namely “Chingola road”, “Nkana East”, “Chisokone”, “Zamtan”, “Chamboli”, and “Naka yombo”. These were the only council mortality data capture centres servicing each entire district respectively.

### Estimation of Excess Mortality

For each month, the pre-pandemic ACM data (up to December 2019) was used to estimate the average expected ACM for the corresponding month during the COVID-19 pandemic period (January 2020 to December 2021) using both Simple Linear Regression and Negative Binomial Regression methods. From this, Excess mortality for each month during the pandemic was then estimated using the following formula:

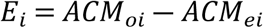

Where *E_i_* is the estimated excess mortality for month *i*, *ACM_oi_* is the total observed all-cause mortality (total burial permits) for month *i*, and *ACM_ei_* is the regression estimated average expected all-cause mortality for month *i* during the COVID-19 pandemic. When using Simple Linear Regression, *ACM_ei_* was estimated as follows:

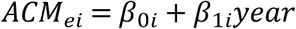

Where *β*_0*i*_ and *β*_1*i*_ are the regression intercept and coefficient for month *i* respectively and “year” is the independent variable. Simple Linear Regression is an Ordinary Least Squares (OLS) regression technique that predicts the mean expected value of a discrete outcome, in this case *ACM_ei_* [50,51]. On the other hand, when using Negative Binomial Regression, *ACE_ei_* was estimated as follows:

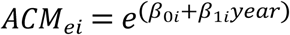

where *β*_0*i*_ and *β*_1*i*_ are the regression intercept and coefficient for month *i* respectively representing the change in the log count for a one-unit change in the independent variable, “year” [52,53]. Negative binomial regression is a method used to forecast outcomes that are counts taking place within a specific area or time frame – in this case a month [54,55,56,57]. In contrast to OLS regression, Negative binomial regression does not assume a linear relationship between the independent and dependent variable and does not require the residuals to be normally distributed with a constant variance which is a requirement in OLS regression [56,57,58]. This is useful especially when working with small datasets.

### Correlation between Excess Mortality, COVID-19 Incidence, and Meteorological Parameters

Pearson’s correlation was used to assess the relationship between the estimated COVID-19 associated excess mortality and COVID-19 incidence for all districts. Further, Pearson’s correlation and multivariate linear regression were also used to explore the relation between the estimated COVID-19 associated excess mortality and average monthly temperature, average monthly relative humidity, and average monthly hours of sunshine using data from Lusaka districts.

### Data collection permits and data analysis tools

All data were collected, cleaned, and summarised in Microsoft Excel software 2021 [59,60]. All statistical data analyses were performed in IBM SPSS software version 20 [61]. Authority to conduct the research was obtained from the Zambia National Health Research Authority NHRA (Ref No: NHRA000047/30/03/2022). Ethical approval for this study was obtained from the University of Zambia Biomedical Research Ethics Committee (UNZABREC Ref ID No. 2479-2021). A waiver of consent was approved by the ethics committee since the study used secondary burial data. Authority to collect council burial permits data was obtained from the offices of the town clerk for Lusaka, Ndola, and Kitwe districts respectively. Official COVID-19 data for each of the study districts in Zambia was collected from the Zambia National Public Health Institute (ZNPHI) with permission obtained from the office of the permanent secretary of the Ministry of Health of Zambia. Permission to collect official meteorological data for the Lusaka district was obtained from the office of the director of the Zambia Meteorological Department in Lusaka where the meteorological data was collected. Finaly, authority to publish and disseminate study findings was granted by the Zambia National Health Research Authority NHRA (**S1** File).

### Data gaps

Data gaps due to torn or missing pages in the burial registers were filled by tracking the serial numbers between the gaps. This process was done to complete the death counts for October 2020 in Kitwe district and July 2019 in Ndola district for which data was partly missing. However, data for the entire year of 2017 were missing for Kitwe district and so this year was completely omitted from the regression analysis. Similarly, for the year 2015 in the Lusaka district, only data from May to August were available. Therefore, only these months were included in the regression analysis from the year 2015 for the district.

## Results

### Unavailable mortality data

The Lusaka District Council had burial permit data for all the major burial sites. However, over 40 other minor traditional burial sites were identified in the Ndola district for which the Ndola city council had no mortality data. Similarly, about 8 other traditional burial sites were identified in Kitwe district for which the Kitwe city council had no mortality data.

### Total all-cause mortalities recorded

Based on burial permit records, there were just over 113,000 total mortalities from all causes between 2015 and 2021 in the Lusaka district (January – April 2015 and September – December 2015 data excluded). Of these total mortalities, 37,400 were recorded between 2020 and 2021 during the COVID-19 pandemic period. On the other hand, about 50,900 total mortalities were recorded between 2011 and 2021 in the Ndola district of which 7,173 occurred between 2020 and 2021 during the COVID-19 pandemic. Kitwe district had just about 28,100 total mortalities between 2015 and 2021 (2017 data excluded) and of this, 7,328 were recorded between 2020 and 2021 during the COVID-19 pandemic.

### Estimated monthly average expected all-cause mortalities (ACM) using linear regression vs negative binomial regression methods

Tables 1 – 3 show the results of the estimated monthly average expected all-cause mortalities (*ACM_ei_*) in the study period for Lusaka, Ndola, and Kitwe districts using the linear regression and the negative binomial regression methods. The binomial regression method consistently estimated *ACM_ei_* values that were either equal to or slightly higher than those estimated by linear regression. The reason for this was unclear apart from the difference in estimation methods. However, the absolute difference in the estimated *ACM_ei_*values for each month between the two methods was very minimal. For example, the largest observed difference in the estimated *ACM_ei_* values between the two methods in Lusaka district was only a 0.91% difference for May 2021 while in Ndola it was a 5.43 % difference for October 2021 and a 10.26 % difference for July 2021 in Kitwe (Tables 1 – 3). Similarly, the difference in the cumulative estimated *ACM_ei_* values over the entire study period between the two methods were only 0.22% for Lusaka, 2.06% for Ndola, and only 1.81% for Kitwe district (last row in Tables 1 – 3). Since the results were generally consistent between the two methods, excess mortality for each month is reported based on the *ACM_ei_* baseline estimated through the linear regression method only for the rest of this study. This is because linear regression is a more stringent estimation method and was able to provide slightly conservative estimates of the excess mortality since its estimated *ACM_ei_* values were generally equal to or slightly lower than those estimated by binomial regression. This way, binomial regression only served as a supplementary method to independently verify the *ACM_ei_* values derived from the linear regression analysis.

**Table 1.** Comparison of the estimated monthly average expected all-cause mortalities (*ACM_ei_*) between the linear regression and the negative binomial regression methods for Lusaka district.

| Lusaka |  |  |  |
| --- | --- | --- | --- |
| | Linear regression | Binomial regression | % difference in estimated $ACM_{ei}$<br>between the two methods. |
| Month | $ACM_{ei}$ | $ACM_{ei}$ | |
| Mar 2020 | 1328 | 1327 | 0.08% |
| Apr 2020 | 1294 | 1297 | 0.27% |
| May 2020 | 1319 | 1325 | 0.49% |
| Jun 2020 | 1400 | 1400 | 0.01% |
| Jul 2020 | 1515 | 1515 | 0.01% |
| Aug 2020 | 1429 | 1429 | 0.01% |
| Sep 2020 | 1450 | 1455 | 0.34% |
| Oct 2020 | 1647 | 1650 | 0.18% |
| Nov 2020 | 1518 | 1518 | 0.03% |
| Dec 2020 | 1477 | 1479 | 0.14% |
| Jan 2021 | 1363 | 1362 | 0.08% |
| Feb 2021 | 1234 | 1239 | 0.41% |
| Mar 2021 | 1275 | 1278 | 0.20% |
| Apr 2021 | 1229 | 1239 | 0.81% |
| May 2021 | 1271 | 1283 | 0.91% |
| Jun 2021 | 1395 | 1395 | 0.01% |
| Jul 2021 | 1504 | 1503 | 0.05% |
| Aug 2021 | 1424 | 1424 | 0.01% |
| Sep 2021 | 1484 | 1494 | 0.67% |
| Oct 2021 | 1687 | 1693 | 0.38% |
| Nov 2021 | 1535 | 1537 | 0.14% |
| Dec 2021 | 1493 | 1496 | 0.17% |
| **Total** | **31270** | **31338** | **0.22%** |

**Table 2.** Comparison of the estimated monthly average expected all-cause mortalities (*ACM_ei_*) between the linear regression and the negative binomial regression methods for Ndola district.

| Ndola |  |  |  |
| --- | --- | --- | --- |
| | Linear regression | Binomial regression | % difference in estimated $ACM_{ei}$<br>between the two methods. |
| Month | $ACM_{ei}$ | $ACM_{ei}$ | |
| Mar 2020 | 311 | 312 | 0.21% |
| Apr 2020 | 300 | 301 | 0.45% |
| May 2020 | 309 | 313 | 1.29% |
| Jun 2020 | 282 | 287 | 1.84% |
| Jul 2020 | 311 | 313 | 0.79% |
| Aug 2020 | 324 | 328 | 1.18% |
| Sep 2020 | 269 | 270 | 0.35% |
| Oct 2020 | 287 | 294 | 2.40% |
| Nov 2020 | 273 | 277 | 1.47% |
| Dec 2020 | 281 | 285 | 1.50% |
| Jan 2021 | 305 | 309 | 1.31% |
| Feb 2021 | 275 | 281 | 2.09% |
| Mar 2021 | 288 | 293 | 1.66% |
| Apr 2021 | 280 | 286 | 2.15% |
| May 2021 | 292 | 300 | 2.57% |
| Jun 2021 | 259 | 270 | 4.44% |
| Jul 2021 | 296 | 300 | 1.44% |
| Aug 2021 | 305 | 313 | 2.62% |
| Sep 2021 | 244 | 251 | 2.78% |
| Oct 2021 | 261 | 275 | 5.43% |
| Nov 2021 | 249 | 259 | 4.18% |
| Dec 2021 | 251 | 263 | 4.90% |
| **Total** | **6251** | **6380** | **2.06%** |

**Table 3.** Comparison of the estimated monthly average expected all-cause mortalities (*ACM_ei_*) between the linear regression and the negative binomial regression methods for Kitwe district.

| Kitwe |  |  |  |
| --- | --- | --- | --- |
| | Linear regression | Binomial regression | % difference in estimated $ACM_{ei}$<br>between the two methods. |
| Month | $ACM_{ei}$ | $ACM_{ei}$ | |
| Mar 2020 | 381 | 382 | 0.26% |
| Apr 2020 | 403 | 403 | 0.00% |
| May 2020 | 342 | 347 | 1.46% |
| Jun 2020 | 337 | 338 | 0.30% |
| Jul 2020 | 347 | 365 | 5.19% |
| Aug 2020 | 406 | 407 | 0.25% |
| Sep 2020 | 311 | 317 | 1.93% |
| Oct 2020 | 410 | 411 | 0.24% |
| Nov 2020 | 402 | 403 | 0.25% |
| Dec 2020 | 374 | 387 | 3.48% |
| Jan 2021 | 378 | 383 | 1.32% |
| Feb 2021 | 352 | 356 | 1.14% |
| Mar 2021 | 364 | 366 | 0.55% |
| Apr 2021 | 402 | 402 | 0.00% |
| May 2021 | 313 | 324 | 3.51% |
| Jun 2021 | 313 | 318 | 1.60% |
| Jul 2021 | 302 | 333 | 10.26% |
| Aug 2021 | 395 | 397 | 0.51% |
| Sep 2021 | 280 | 293 | 4.64% |
| Oct 2021 | 398 | 399 | 0.25% |
| Nov 2021 | 395 | 396 | 0.25% |
| Dec 2021 | 340 | 362 | 6.47% |
| **Total** | **7945** | **8089** | **1.81%** |

### Estimated monthly excess mortalities

Lusaka district had an 11.1% overall increase in expected total deaths (3,484 excess mortalities) between March 2020 – December 2021 during the COVID-19 pandemic (Figure 1). Significant excess mortalities were observed, especially in the months of June and July 2021. Ndola district recorded an estimated 378 excess mortalities which was a 6.1% increase overall in total expected deaths in the same study period (March 2020 – December 2021) during the COVID-19 pandemic (Figure 2). In Ndola, Significant excess mortalities were also observed, especially in the months of June and July 2021. On the other hand, Kitwe district actually recorded 16.9% fewer deaths than expected overall between March 2020 – December 2021 during the COVID-19 pandemic (Figure 3). Despite this, some isolated months (October 2020 as well as June 2021 and July 2021) did record excess mortalities in Kitwe (Figure 5A, 5B) with significant excess deaths recorded in October 2020.

**Figure 1.**
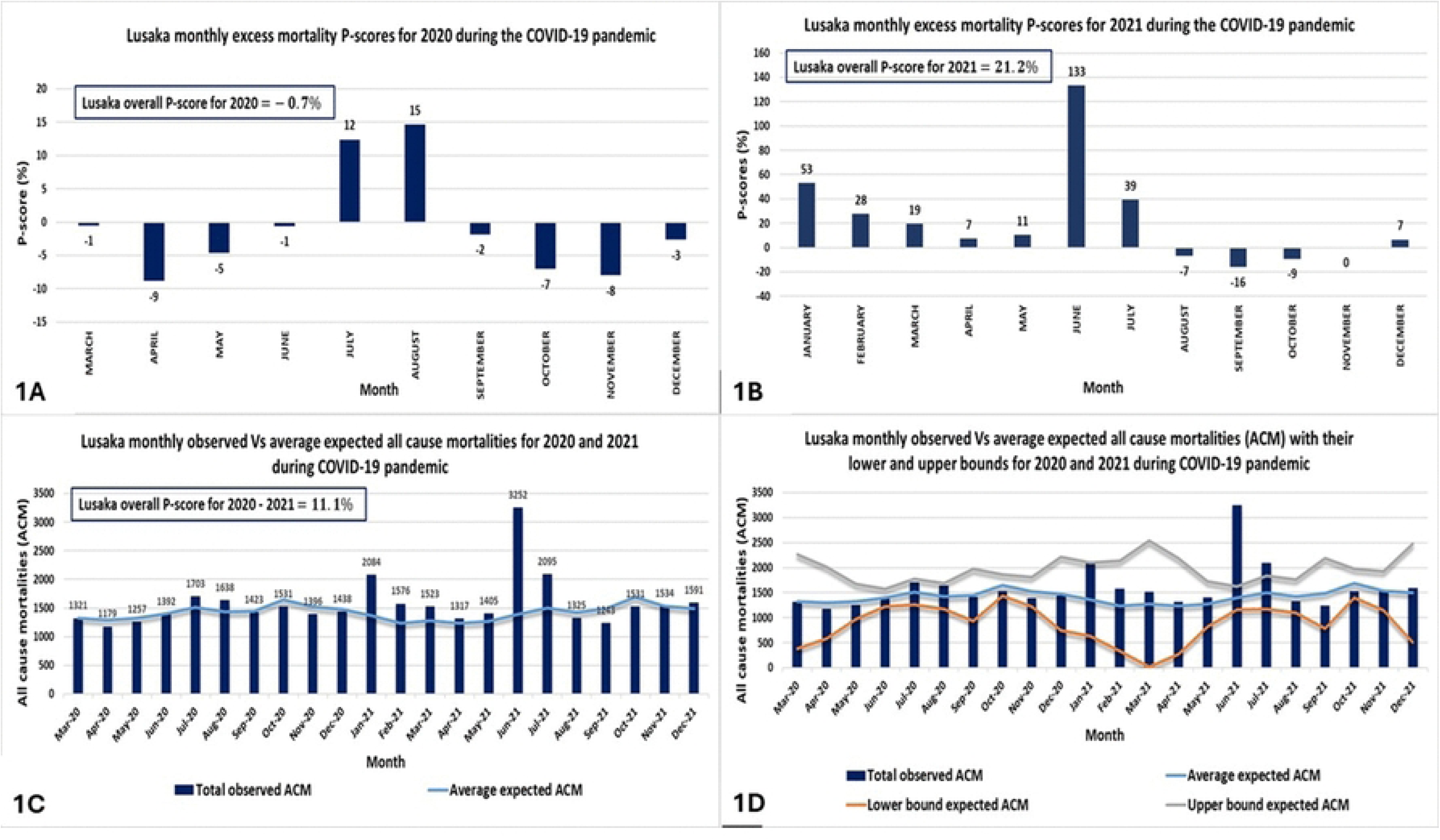
Lusaka - monthly excess mortality p-scores for **(1a)** 2020 and **(1b)** 2021. **(1c)** observed monthly all cause mortalities (ACM) for 2020 – 2021 vs monthly average expected ACM, **(1d)** 2020 – 2021 estimated monthly average expected ACM confidence intervals.

**Figure 2.**
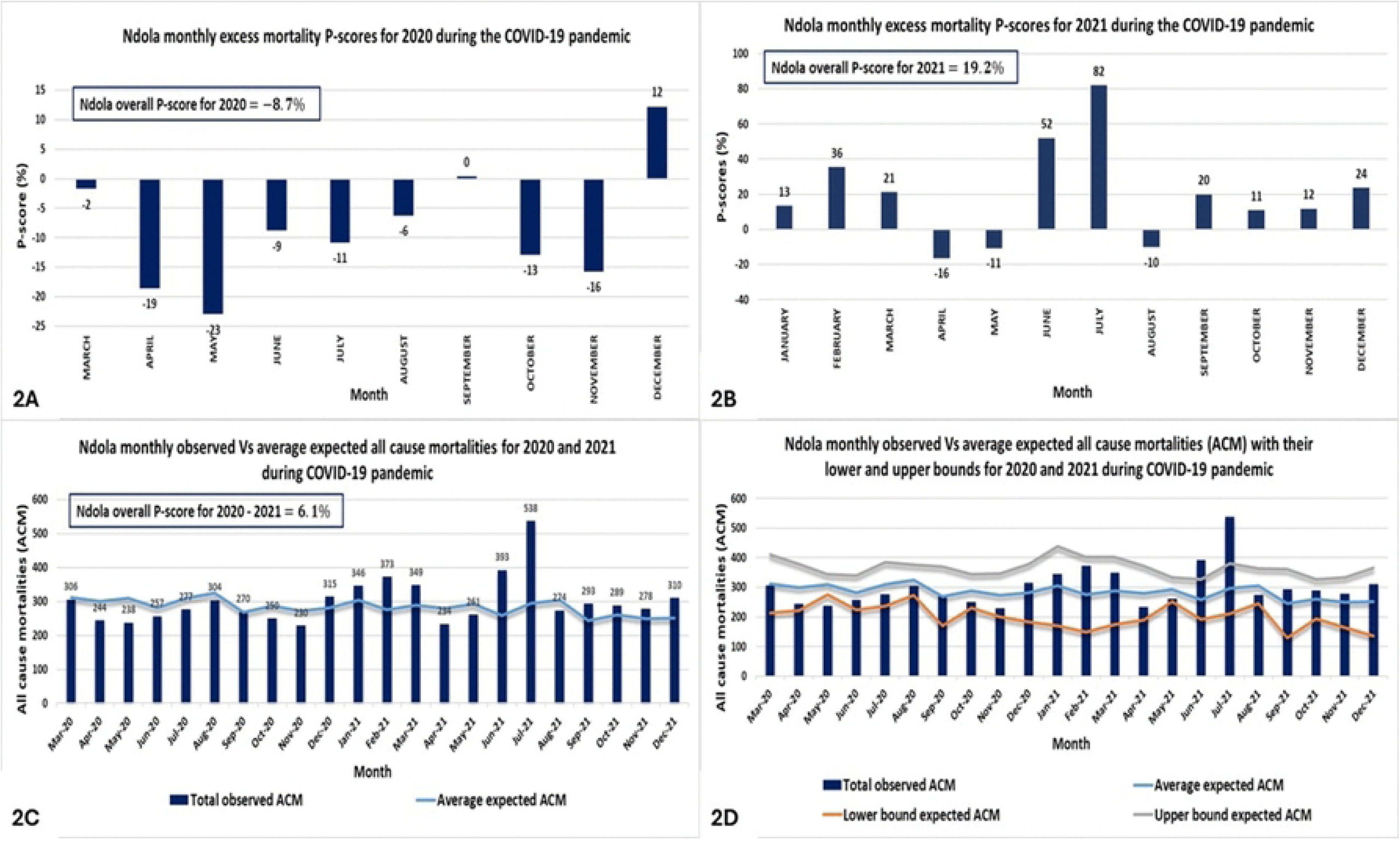
Ndola - monthly excess mortality p-scores for **(2a)** 2020 and **(2b)** 2021. **(2c)** observed monthly all cause mortalities (ACM) for 2020 – 2021 vs monthly average expected ACM, **(2d)** 2020 – 2021 estimated monthly average expected ACM confidence intervals.

**Figure 3.**
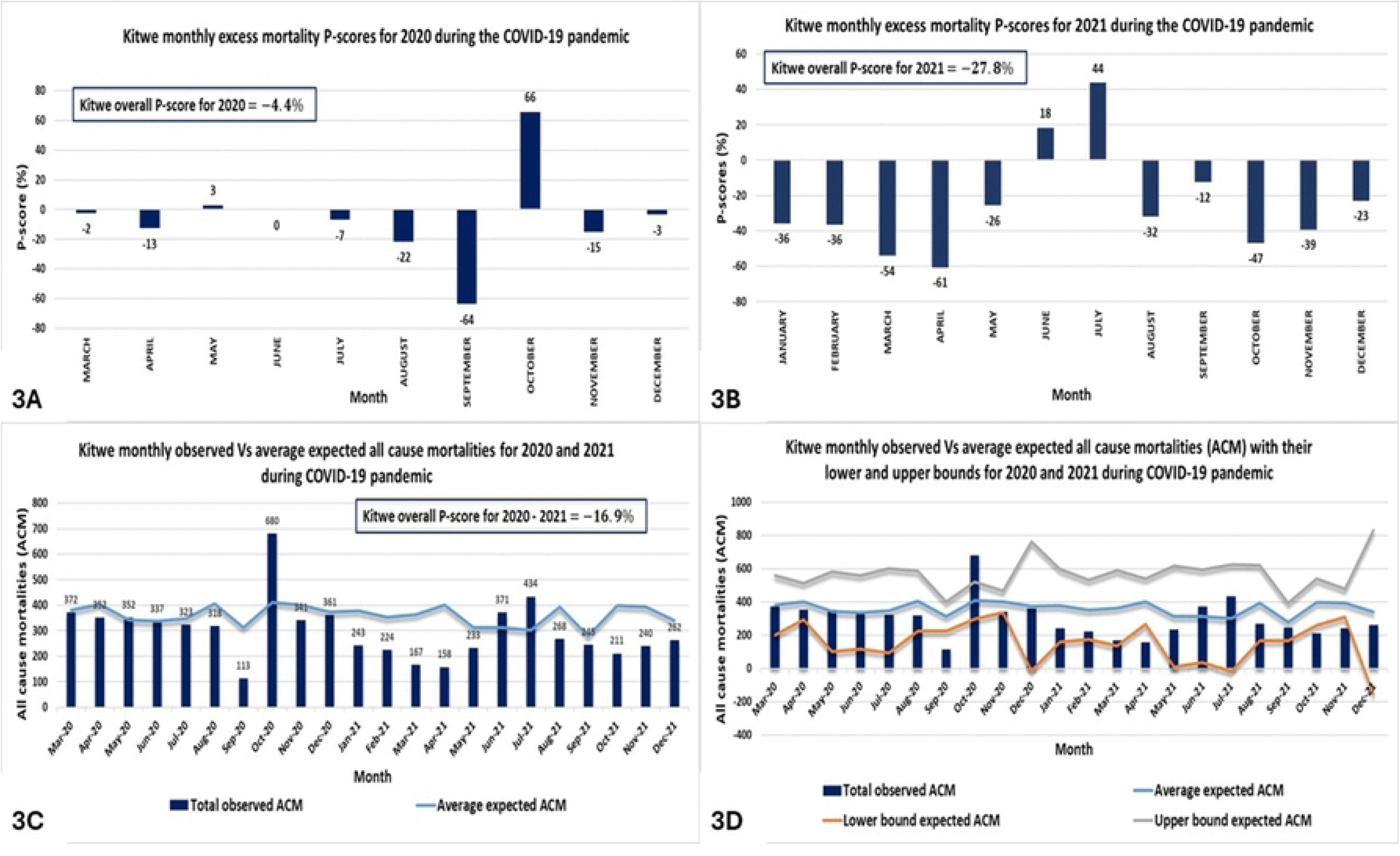
Kitwe - monthly excess mortality p-scores for **(3a)** 2020 and **(3b)** 2021. **(3c)** observed monthly all cause mortalities (ACM) for 2020 – 2021 vs monthly average expected ACM, **(3d)** 2020 – 2021 estimated monthly average expected ACM confidence intervals.

### Correlation between excess mortalities and COVID-19 incidence and deaths

The estimated monthly excess mortalities in this study had a significant positive correlation with the officially reported monthly COVID-19 incidence and deaths for both Lusaka and Ndola districts respectively (P < 0.05, Figures 4 and 5). However, a positive but non-significant correlation was observed in Kitwe district (Figure 6).

**Figure 4.**
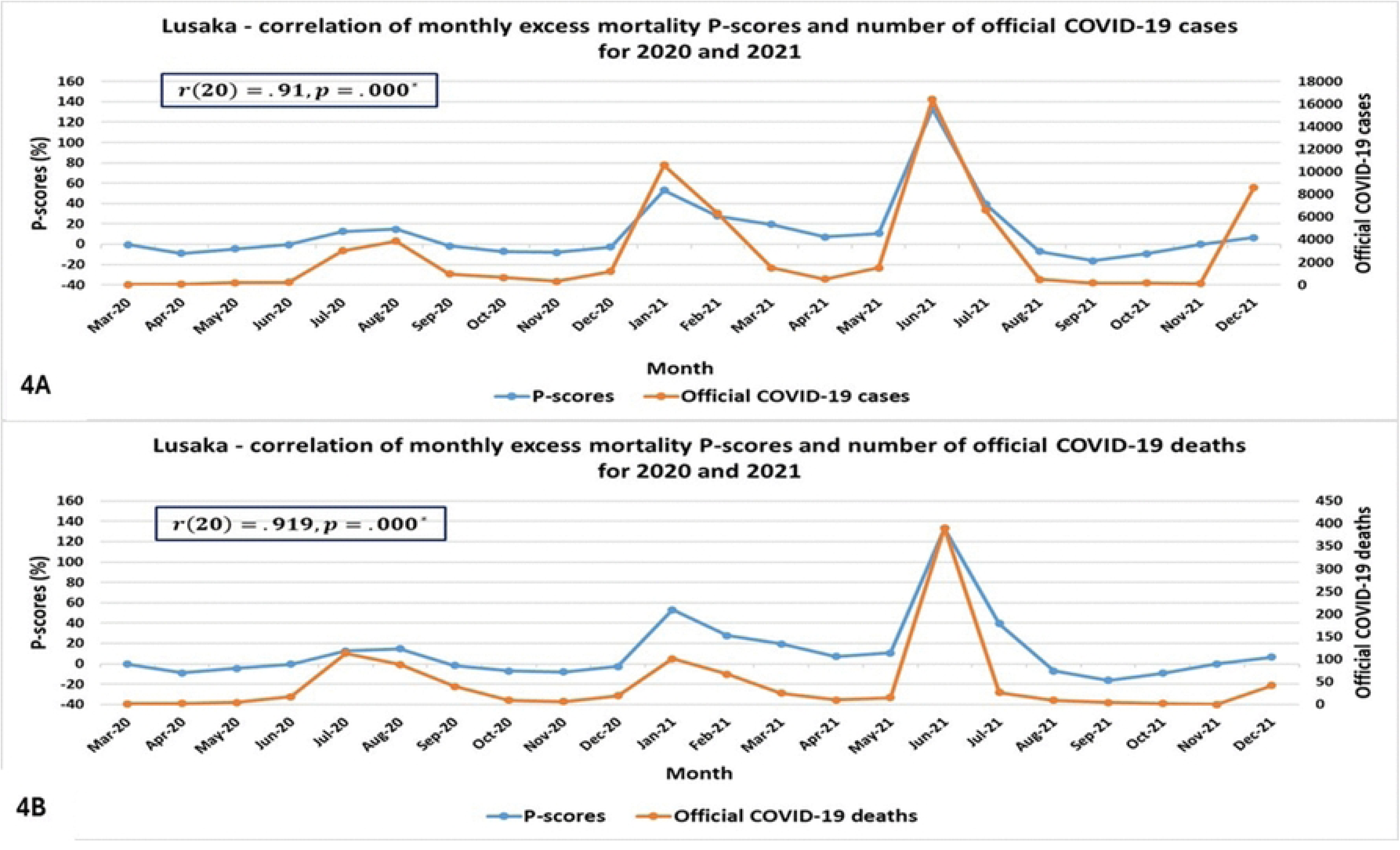
Lusaka - correlation of monthly excess mortality P-scores with **(4a)** official COVID-19 cases for 2020 – 2021, **(4b)** official COVID-19 deaths for 2020 – 2021.

**Figure 5.**
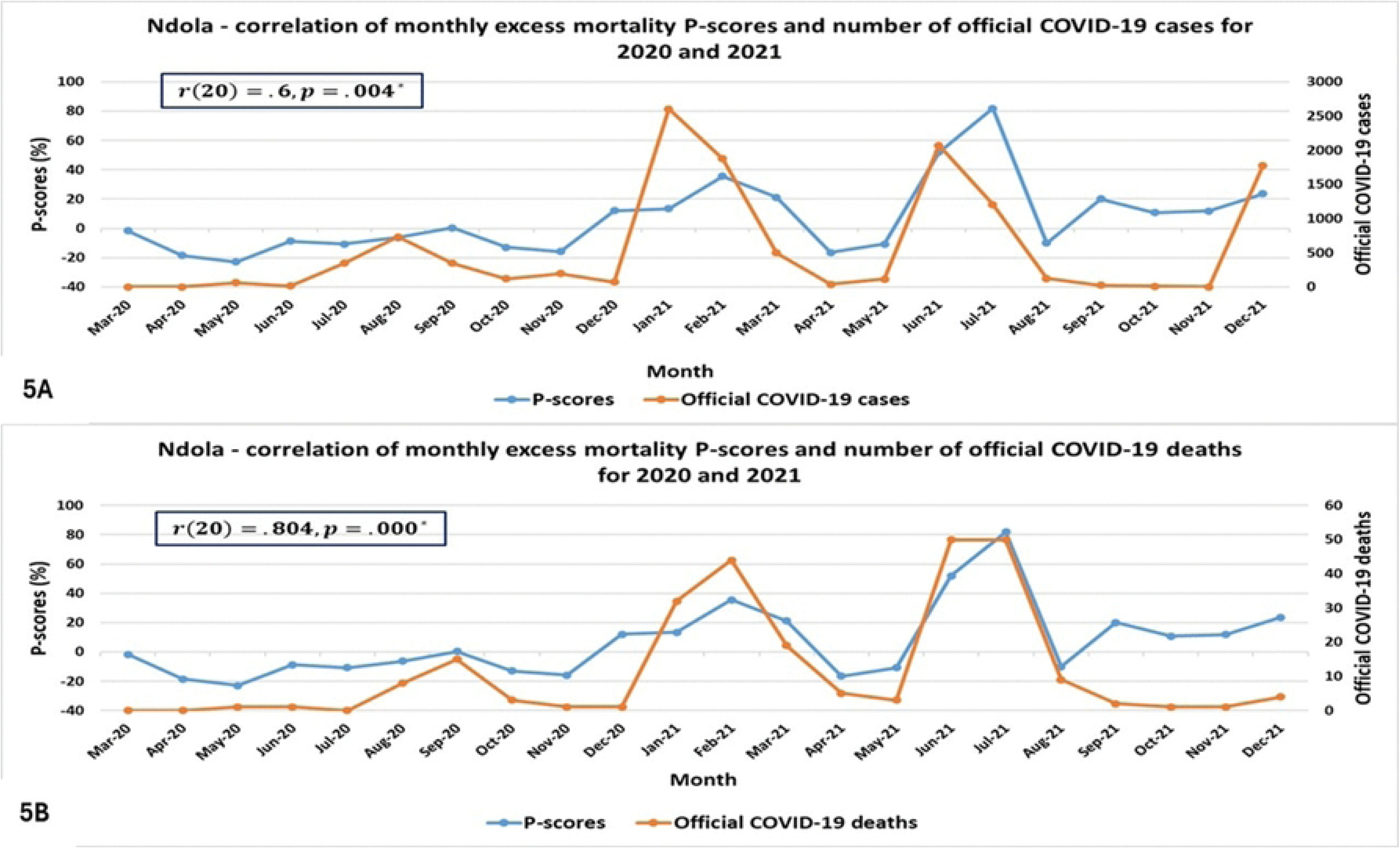
Ndola - correlation of monthly excess mortality P-scores with **(5a)** official COVID-19 cases for 2020 – 2021, **(5b)** official COVID-19 deaths for 2020 – 2021.

**Figure 6.**
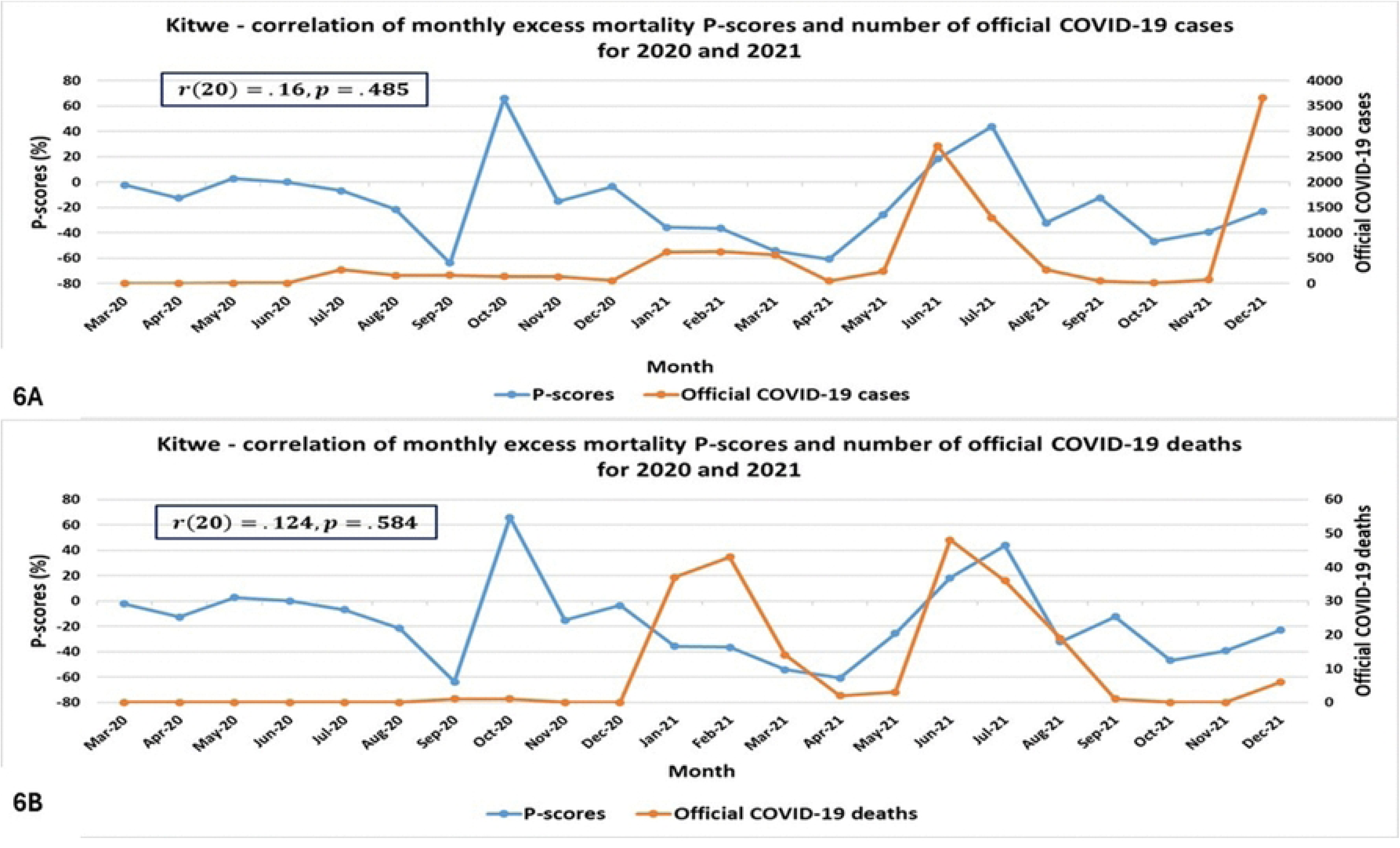
Kitwe - correlation of monthly excess mortality P-scores with **(6a)** official COVID-19 cases for 2020 – 2021, **(6b)** official COVID-19 deaths for 2020 – 2021.

### Correlation between excess mortality and meteorological parameters in Lusaka district

In this study, excess mortality generally negatively correlated with temperature with a significant negative correlation observed between excess mortality and the average monthly maximum temperature (Figure 7). Further, it was also observed that the ambient temperature; either as the average monthly minimum, mean, or maximum temperature, was a significant predictor of both COVID-19 incidence as well as COVID-19 related excess mortality after controlling for the other meteorological parameters namely the average monthly relative humidity and the average monthly hours of sunshine based on the multivariate linear regression analysis (Tables 4 – 6 and Tables 7 – 9). In fact, ambient temperature more consistently predicted excess mortality than COVID-19 incidence after controlling for the other metrological parameters (Tables 4 – 6 and Tables 7 – 9).

**Figure 7.**
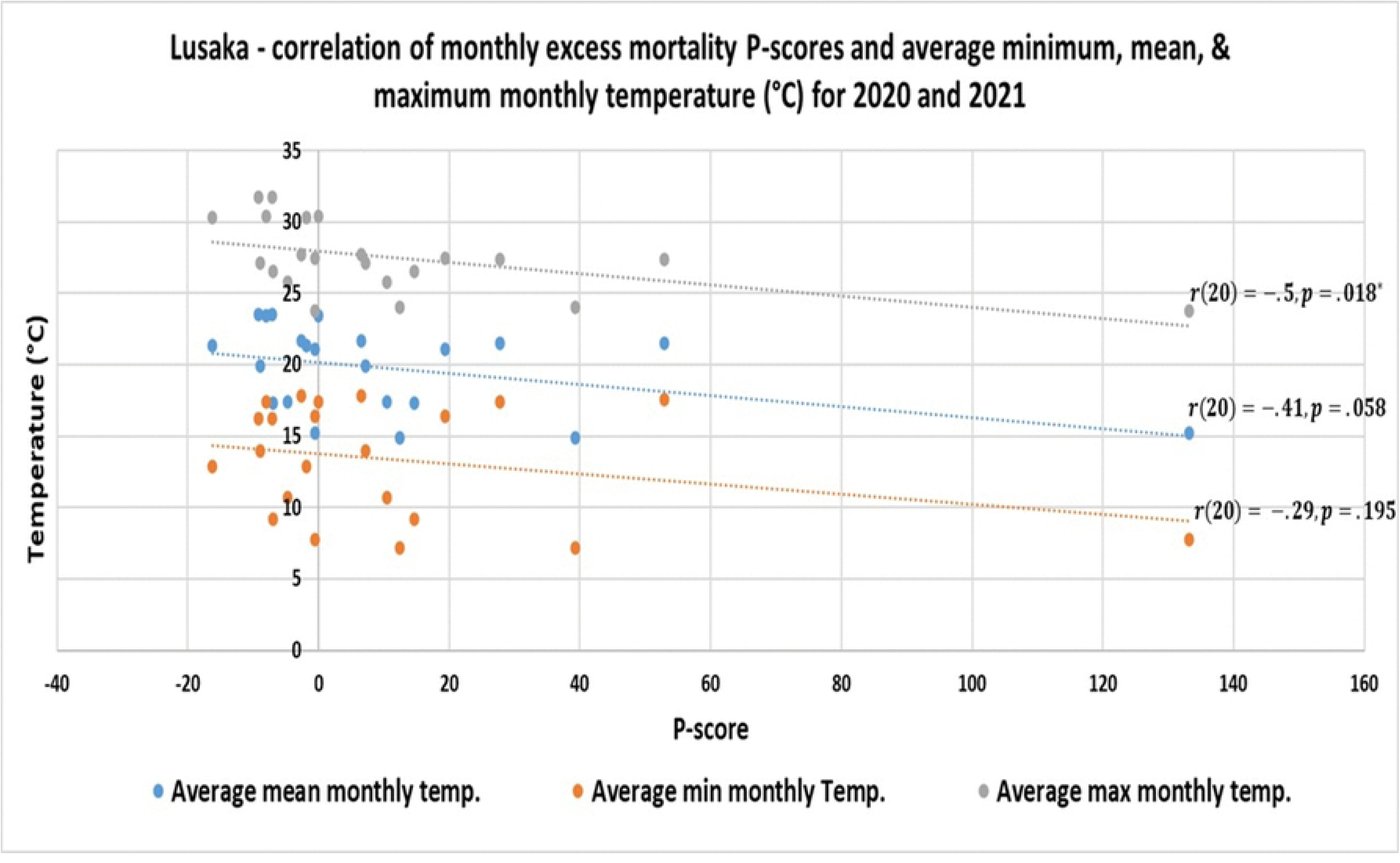
Lusaka - correlation of monthly excess mortality P-scores and average monthly minimum, mean, and maximum temperatures (°C) for 2020 – 2021)

**Table 4.** Average monthly mean temperature (°C), average monthly relative humidity, and average monthly hours of sunshine as predictors of excess mortality in Lusaka district during the pandemic.

| **<u>Model Summary; Multivariate Linear Regression for Lusaka Data:</u>** |  |  |
| --- | --- | --- |
| *Overall model significance test: ANOVA / Sig. F Change (P-value) = .046\** |  |  |
| **<u>Dependent Variable</u>** | **<u>Predictors</u>** |  |
| Monthly excess mortality (P-score) | Average monthly **mean** temperature<br>Average monthly relative |  |
| ***Predictors*** | ***Unstandardized coefficients (95% CI)*** | ***(P-value)*** |
| (Constant) | 493.439 (26.473 → 960.406) | .053 |
| Average monthly mean temperature | -10.012 (-17.089 → -2.934) | **.013\*** |
| Average monthly relative humidity | 1.197 (-3.777 → 0.915) | .247 |
| Average monthly hours of sunshine | -.757 (-1.552 → 0.037) | .078 |

\*Significant at 0.05 level of significance.

**Table 5.** Average monthly maximum (max) temperature (°C), average monthly relative humidity, and average monthly hours of sunshine as predictors of excess mortality in Lusaka district during the pandemic.

| **<u>Model Summary; Multivariate Linear Regression for Lusaka Data:</u>** |  |  |
| --- | --- | --- |
| *Overall model significance test: ANOVA / Sig. F Change (P-value) = .045\** |  |  |
| **<u>Dependent Variable</u>**<br>Monthly excess mortality (P-score) |  | **<u>Predictors</u>**<br>Average monthly **max** temperature<br>Average monthly relative |
| ***Predictors*** | ***Unstandardized coefficients (95% CI)*** | ***(P-value)*** |
| (Constant) | 611.075 (68.800 → 1153.349) | .040 |
| Average monthly max temperature | -11.688 (-19.936 → -3.440) | **.012\*** |
| Average monthly relative humidity | -1.922 (-4.536 → 0.692) | .167 |
| Average monthly hours of sunshine | -.606 (-1.313 → 0.101) | .110 |

\*Significant at 0.05 level of significance.

**Table 6.** Average monthly minimum (min) temperature (°C), average monthly relative humidity, and average monthly hours of sunshine as predictors of excess mortality in Lusaka district during the pandemic.

| **<u>Model Summary; Multivariate Linear Regression for Lusaka Data:</u>** |  |  |
| --- | --- | --- |
| *Overall model significance test: ANOVA / Sig. F Change (P-value) = .036\** |  |  |
| **<u>Dependent Variable</u>** | **<u>Predictors</u>** |  |
| Monthly excess mortality (P-score) | Average monthly **min** temperature<br>Average monthly relative humidity |  |
| ***Predictors*** | ***Unstandardized coefficients (95% CI)*** | ***(P-value)*** |
| (Constant) | 433.560 (20.400 → 846.721) | .054 |
| Average monthly min temperature | -9.343 (-15.657 → -3.029) | **.010\*** |
| Average monthly relative humidity | -.918 (-2.947 → 1.111) | .387 |
| Average monthly hours of sunshine | -.942 (-1.815 → -0.070) | **.048\*** |

\*Significant at 0.05 level of significance.

**Table 7.** Average monthly mean temperature (°C), average monthly relative humidity, and average monthly hours of sunshine as predictors of confirmed COVID-19 cases (incidence) in Lusaka district during the pandemic.

|  |  |
| --- | --- |
| Official confirmed COVID-19 cases | Average monthly **mean** temperature<br>Average monthly relative |

| ***Predictors*** | ***Unstandardized coefficients (95% CI)*** | ***(P-value)*** |
| --- | --- | --- |
| (Constant) | 96232.131 (39084.584 → 153379.678) | .004 |
| Average monthly mean temperature | -1599.939 (-2466.112 → -733.765) | **.002\*** |
| Average monthly relative humidity | -334.016 (-621.095 → -46.937) | **.035\*** |
| Average monthly hours of sunshine | -158.572 (-255.791 → -61.354) | **.005\*** |

\*Significant at 0.05 level of significance.

**Table 8.** Average monthly maximum (max) temperature (°C), average monthly relative humidity, and average monthly hours of sunshine as predictors of confirmed COVID-19 (incidence) cases in Lusaka district during the pandemic.

| **<u>Model Summary; Multivariate Linear Regression for Lusaka Data:</u>** |  |  |
| --- | --- | --- |
| *Overall model significance test: ANOVA / Sig. F Change (P-value) = .010\** |  |  |
| **<u>Dependent Variable</u>**<br>Official confirmed COVID-19 cases |  | **<u>Predictors</u>**<br>Average monthly **max** temperature<br>Average monthly relative humidity |
| ***Predictors*** | ***Unstandardized coefficients (95% CI)*** | ***(P-value)*** |
| (Constant) | 113728.475 (46841.218 → 180615.732) | .004 |
| Average monthly max temperature | -1846.504 (-2863.852 → -829.156) | **.002\*** |
| Average monthly relative humidity | -406.987 (-729.381 → -84.593) | **.024\*** |
| Average monthly hours of sunshine | -132.981 (-220.237 → -45.725) | **.008\*** |

\*Significant at 0.05 level of significance.

**Table 9.** Average monthly minimum (min) temperature (°C), average monthly relative humidity, and average monthly hours of sunshine as predictors of confirmed COVID-19 (incidence) cases in Lusaka district during the pandemic.

| **<u>Model Summary; Multivariate Linear Regression for Lusaka Data:</u>** |  |  |
| --- | --- | --- |
| *Overall model significance test: ANOVA / Sig. F Change (P-value) = .009\** |  |  |
| **<u>Dependent Variable</u>**<br>Official confirmed COVID-19 cases |  | **<u>Predictors</u>**<br>Average monthly **min** temperature<br>Average monthly relative humidity |
| ***Predictors*** | ***Unstandardized coefficients (95% CI)*** | ***(P-value)*** |
| (Constant) | 84406.548 (33302.371 → 117708.919) | .005 |
| Average monthly min temperature | -1453.751 (-2234.723 → -672.778) | **.002\*** |
| Average monthly relative humidity | -243.664 (-494.606 → 7.278) | .073 |
| Average monthly hours of sunshine | -183.468 (-291.401 → -75.534) | **.004\*** |

\*Significant at 0.05 level of significance.

## Discussion

This study aimed to estimate excess mortality associated with the COVID-19 pandemic using burial records data in three highly impacted major districts in Zambia namely Lusaka, Ndola, and Kitwe districts. It was estimated in this study that Lusaka district recorded about 3,484 excess mortalities (11.1% increase in total expected deaths) overall between March 2020 and December 2021 during the COVID-19 pandemic (Figure 1c). Assuming that all excess mortalities were associated with COVID-19, this means that during this period, COVID-19 associated deaths were approximately 3.5 times more than what was officially reported through laboratory confirmed COVID-19 deaths in Lusaka district. Most of these excess mortalities occurred in the year 2021 where 21.2% excess deaths were recorded between January 2021 and December 2021 compared to what was expected for that period alone in Lusaka (Figure 1b). Because COVID-19 is postulated to have caused the excess mortality in this study, the high excess mortality in 2021 was likely due to the two largest waves of the COVID-19 pandemic in Zambia that both occurred in 2021 (the second large wave between January 2021 to March 2021 and the third and largest wave between June 2021 to August 2021) [62]. These waves may have also been fuelled by the circulation of the highly transmissible and virulent Delta variant of the SARS-CoV-2 virus (the causative agent for COVID-19) which dominated for much of 2021 globally [63,64]. Additionally, despite the COVID-19 vaccination program being officially launched in April 2021 in Zambia, The Zambian population exhibited high vaccine hesitancy and low vaccine uptake [65, 66, 67]. For example, by mid-August 2021 near the end of the third COVID-19 wave, only about 1.5% of the Zambian population had received at least one dose of a COVID-19 vaccine [68, 69]. This likely contributed to more serious illness and undiagnosed COVID-19 deaths in the community if we also consider the low testing rates in the country [11,18, 43]. The results for Lusaka are consistent with the estimates by Msemburi and colleagues who estimated that Zambia recorded between 10% to 20% excess mortalities during the COVID-19 pandemic from January 2020 to December 2021 since Lusaka was one of the major contributing districts. Ndola and Kitwe districts data was considered incomplete for accurate estimation of excess mortality in this study due to gross data gaps owing to the presence of numerous traditional burial sites in these districts for which the respective city councils had insufficient data. Therefore, results for Ndola and Kitwe were interpretated cautiously. notwithstanding, excess mortalities were still detected in Ndola district where about 378 excess deaths (6.1% increase in overall expected mortalities) were recorded in the district between March 2020 – December 2021 (Figure 2c). Again, assuming COVID-19 was linked to all excess mortalities, this translates into 1.5 times more COVID-19 associated deaths than what was officially reported through laboratory confirmed COVID-19 deaths in Ndola district. Like Lusaka, most of these excess mortalities in Ndola occurred in 2021 where 19.2% more deaths than expected were recorded between January 2021 and December 2021 probably for the same reasons discussed above (Figure 2b). It is possible that the missing mortality data in Ndola could have affected this result, and the actual excess mortality may have been higher in the district overall. Kitwe district on the other hand recorded 16.9% fewer deaths overall than what was expected between March 2020 – December 2021 (Figure 3c) despite recording excess mortalities in the isolated months of October 2020 as well as June 2021 and July 2021 in the district (Figures 3a – 3c). A drop in total expected all cause mortalities during the COVID-19 pandemic can result for example from instituted stringent infection prevention measures and lock-downs causing a drop in deaths from other natural infectious diseases in the population [16]. This effect was observed in countries like Australia, New Zealand, and Uruguay [16, 70]. However, Like Ndola district, because of the missing mortality data in Kitwe, it was inconclusive as to whether this was a genuine overall drop in total expected mortalities during this period in the district or a result of data bias.

An accurate estimation of the average expected all-cause mortalities (ACL) in a particular place and time is an important pre-requisite for estimating excess mortality. This parameter reflects the natural rate of all cause deaths in the population determined by stable factors such as the age composition of the population, the incidence of smoking and air pollution, the prevalence of obesity, poverty and inequality, and the normal quality of health service delivery among others [71]. There was generally very strong agreement in the estimated monthly average expected all cause mortalities (*ACM_ei_* ) between the linear regression and the negative binomial regression methods for all the districts during the pandemic as seen in tables 1 (Lusaka), 2 (Ndola), and 3 (Kitwe) in the results section. Overall, this was considered a sufficient level of agreement and strong evidence of high accuracy in the estimated monthly *ACM_ei_* values for each month due to the use of two independent methods to estimate the same parameter. The strongest agreement however was observed in Lusaka district where there was no more than 0.91% difference in the estimated *ACM_ei_* for each month between the two methods.

Interpreted in terms of the outcome variable, the largest difference in the estimated monthly excess mortality between the two methods in Lusaka district was in May 2021 where there is an estimated excess mortality of 9.5% for the month based on the binomial regression *ACM_ei_* baseline (not displayed in results section) as compared to an excess mortality of 11% based on the Linear regression *ACM_ei_* baseline as reported in the results section. Similarly, the largest gap in the estimated excess mortality between the two methods in Ndola district was in October 2021 with an estimated excess mortality of 5.1% for the month based on the binomial regression *ACM_ei_* baseline (not displayed in results section) as compared to the excess mortality of 11% based on the Linear regression *ACM_ei_* baseline as shown in Table 2. Finaly, in Kitwe district the largest difference is observed in July 2021 with an estimated excess mortality of 30% for the month based on the binomial regression *ACM_ei_* baseline (not displayed in results section) as compared to an excess mortality of 44% using the Linear regression *ACM_ei_* baseline as shown in Table 3. The reason for the higher level of consistency in the estimated *ACM_ei_* baseline values, and consequently the monthly excess mortality, between the two methods in Lusaka district as compared to Ndola and Kitwe districts is probably because Lusaka district had more complete mortality data. As earlier mentioned, excess mortality results in this study are reported based on the Linear regression *ACM_ei_* baseline because Linear regression is a more stringent estimation technique. Additionally, the generally slightly lower Linear regression *ACM_ei_* baseline values ensured that conservative estimates of the excess mortality were made for each month to not underestimate this important epidemiological parameter. Lastly, even though the *ACM_ei_* baseline values estimated by linear regression had volatile 95% confidence intervals due to small sample size (Figures 1d, 2d, and 3d), these values were consistent with the average *ACM_ei_* baseline values estimated through the binomial regression which is suitable for small sample sizes as shown in the results section [54,55,56,57, 58].

In Lusaka district, there was a significant positive correlation (P < 0.05) between excess mortality and both the officially confirmed COVID-19 cases and deaths (Figure 4a and 4b). This provided strong supporting evidence that the excess mortality recorded in the district was caused by the COVID-19 pandemic. This is because a rise in officially confirmed COVID-19 cases and deaths, even under weak surveillance systems, usually correlates with increased community infections during a COVID-19 wave. These COVID-19 waves then led to more deaths that were either directly or indirectly associated with COVID-19 including those deaths not captured by the surveillance systems as caused by COVID-19 thereby causing excess mortality. Ndola district data also had a significant positive correlation (P < 0.05) between excess mortalities and officially confirmed COVID-19 cases and deaths (Figure 5a and 5b) while Kitwe district had a positive but non-significant correlation (Figure 6a and 6b). By closely assessing the strength of this correlation and its P-value in each district, the strongest correlation between excess mortalities and officially confirmed COVID-19 cases and deaths is seen in Lusaka district. A possible explanation for this is that the COVID-19 pandemic truly caused the excess mortality as explained by this correlation and the strongest evidence for this is seen in Lusaka district because Lusaka had the most complete mortality data of all the districts in this study. On the other hand, Ndola and Kitwe districts showed slightly weaker positive correlation (although significant in Ndola) because of having relatively incomplete mortality data. Another reason for this could be that since Lusaka was the epicentre of the COVID-19 pandemic in Zambia, it had the highest total incidence of COVID-19 cases and deaths in the country. For example, although Lusaka, Ndola, and Kitwe districts together contributed about 52% (1451) of all confirmed COVID-19 deaths between 2020 and 2021 in Zambia, almost 70% (990) of this 52% was contributed by Lusaka district alone. Similarly, Lusaka district contributed about 26% (63527) of all confirmed COVID-19 cases in Zambia between 2020 and 2021 while Ndola district only contributed about 5% (12228) and Kitwe district only 4.6% (11103) [42,43,72]. This means that out of the three districts, Lusaka district had accumulated enough COVID-19 incidence data in this period to provide more reliable conclusions about the true nature of the dynamics of the pandemic in the population hence more significant correlations observed in Lusaka.

According to various studies [9,16,73] excess mortality during a public health crisis such as the COVID-19 pandemic is effectively the net result of various contributing factors A through E. Factor A are excess deaths directly caused by COVID-19 infection; both laboratory confirmed or unconfirmed. This will depend on the severity of the epidemic in the population. How many of these cases are captured by the national surveillance system will mainly depend on testing capacity and accuracy of attribution of cause of death [9,16]. Factor B are excess deaths caused by collapse of the medical system due to COVID-19 pandemic. This is considered to have minimal effect at least especially in advanced nations [16,17]. In Zambia, this factor also likely had minimal effect if we consider the fact that trends in some important pre-existing public health burdens in the country remained largely unaffected. For example, the Zambian government in collaboration with the Centers for Disease Control and Prevention (CDC) successfully implemented strategies that prevented disruptions to HIV treatment for people living with HIV during the pandemic [74]. Similarly, some reports also found that there were generally no major changes to important child immunization programs in Zambia during the pandemic [75, 76]. Malaria incidence also only rose by a factor of 1.32 [77] while no changes were observed in maternal mortalities overall during the pandemic [78]. Lastly, the WHO also reported that tuberculosis deaths as well as tuberculosis/HIV coinfection deaths continued to drop in Zambia overall even during the pandemic period [76]. Therefore, it is evident that although the health system was strained during the pandemic in Zambia, this alone could not have contributed greatly to the excess mortalities estimated in this study. Factor C are excess deaths from other natural causes (diseases). This factor was observed to have a slight negative effect especially in the context of infectious diseases in other countries [16]. For example, due to stringent lock-down measures implemented in response to COVID-19, countries like Australia, New Zealand, and Uruguay saw a slight drop in winter influenza deaths [16,70]. From the current study, such a phenomenon could have explained the estimated net drop in the overall expected all cause mortalities in Kitwe district. However, the results obtained for Kitwe district were likely affected by incomplete data records and therefore cannot, with certainty, be attributed to this effect as seen in Australia, New Zealand, and Uruguay. Furthermore, to the best of the researcher’s knowledge, no specific reports were found about reductions in other infectious diseases incidence in Zambia as a direct offshoot of the infection prevention measures implemented for COVID-19. Therefore, it is possible this factor did not have a significant effect in Zambia. Factor D are excess deaths due to unnatural causes as influenced by the pandemic such as suicides, homicides, and traffic accidents etc. Again, to the best of the researcher’s knowledge, no data or reports were found about a rise in deaths from these causes during the pandemic in Zambia. Therefore, this factor likely did not have an effect in Zambia. Moreover, the overall effect of this factor during the pandemic contradicted in other countries but was however found to be minimal and did not exceed about 1% of the expected baseline deaths based on data from various studies [16,79,80]. Finally, factor E are excess deaths due to extreme events such as wars and natural disasters etc that may have occurred concurrently within the period of the COVID-19 pandemic. A quick scan of online reports reveals no such major events for Zambia during the pandemic hence this factor can safely be considered to have had no effect. With all these factors discussed above, it can be assumed therefore that most of the excess mortalities estimated for Zambia in this study in the study districts were caused by COVID-19 disease as described by factor A.

Lastly, using Lusaka data only, this study also explored the relation between the estimated COVID-19 related excess mortality and ambient temperature as well as other meteorological parameters. A large volume of epidemiological studies has found that ambient temperature is inversely associated with COVID-19 incidence i.e lower temperatures are associated with higher COVID-19 incidence [81–115]. In Lusaka, it was found that both the estimated excess mortality (Fig 7 and tables 4 – 6) as well as COVID-19 incidence (tables 7 – 8) were inversely associated with the average monthly minimum, mean, or maximum temperature. This means that lower ambient temperatures were associated with higher COVID-19 incidence which led to more excess mortalities in Lusaka. However, it was also observed in this study that generally, ambient temperature is a better single predictor of the COVID-19 related excess mortality (tables 4 – 6) than it is a predictor of COVID-19 incidence (tables 7 – 9) after controlling for the effect of other meteorological parameters namely relative humidity and average monthly hours of sunshine. This is because ambient temperature (either as the average monthly minimum, mean, or maximum temperature) was mostly the only significant predictor in the model for predicting excess mortality (tables 4 – 6) compared to the model for predicting COVID-19 incidence (tables 7 – 9). This is probably because excess mortality is a more objective and unbiased indicator of COVID-19 disease severity and impact on the population as compared to COVID-19 incidence as earlier alluded to and therefore it is more directly sensitive to the effects of ambient temperature related COVID-19 disease severity. Various explanations have been provided for the inverse relationship seen between ambient temperature and COVID-19 disease incidence and severity. For example, the SARS-CoV2 virus, like other respiratory viruses, survives longer in the environment at lower temperatures and low temperatures also likely hinder the human innate immune system because of reduced blood supply in cold weather which reduces provision of immune cells to the nasal mucosa [82,85,88,89,]. The average monthly hours of sunshine also generally negatively correlated with both excess mortality and COVID-19 incidence (tables 4 – 9). This is probably because longer exposure to sunshine is directly proportional to ambient temperature. Relative humidity on the other hand showed both a positive and negative correlation with excess mortality (tables 4 – 9).

## Conclusion

COVID-19 associated deaths were likely 3.5 times higher than what was officially reported in Lusaka district which was the epicentre of the COVID-19 pandemic in Zambia. COVID-19 deaths were also likely higher than what was officially reported in other districts in Zambia as flagged by Ndola district, but this is inconclusive due to incomplete national death registration data or burial records. As seen in Lusaka district, comprehensive local burial records data can be used to track excess mortalities during public health crises in countries lacking complete national death registration data from the national civil registration systems (CRS).

## Data Availability

Data underlying the results presented in the study cannot be shared publicly because it is managed by Zambian government institutions. Permission to collect council burial permits data can be obtained in writing from the offices of the town clerk for Lusaka, Ndola, and Kitwe districts respectively. Permission to collect Official COVID-19 data for each of these study districts can be obtained from the office of the permanent secretary of the Ministry of Health of Zambia. Lastly, permission to collect official meteorological data for Lusaka district can be obtained from the office of the director of the Zambia Meteorological Department in Lusaka where the meteorological data was collected.

## Acknowledgments

I would like to acknowledge Professor Martin Simuunza from the Department of Disease Control, School of Veterinary Medicine, at The University of Zambia for his fundamental role and overall guidance in the implementation of this study. I would also like acknowledge my parents for their continued support throughout my research career.

## Supporting information

**S1 File**. Zambia National Health Research Authority (NHRA) Authority to Publish and Disseminate Study Findings.

